# Real-world Drug Regimens for Multiple Myeloma in a Swiss Population (2012 to 2017): cost-outcome description

**DOI:** 10.1101/19008383

**Authors:** Klaus Eichler, Roland Rapold, Simon Wieser, Oliver Reich, Eva Blozik

## Abstract

**Background:** Novel drugs are dynamically changing current treatment regimens for multiple myeloma (MM). Novel drugs have improved prognosis of MM patients in clinical studies but are expensive. Little is known about up-to-date real-world application and costs.

**Methods:** We performed a retrospective observational cohort analysis (cost-outcome description; 2012-2017) in a claims database of a major Swiss health insurance company which covers 14% of the Swiss population (Helsana Versicherungen AG).We used primary (MM diagnoses via ICD-10) and secondary features (prescribed MM-specific drugs) as inclusion criteria and defined a hierarchy of drug regimens to classify treatments as: 1) proteasome inhibitor (PI)-based regimen (e.g. bortezomib); 2) IMID-based regimen (e.g. lenalidomide); 3) chemotherapy (CHEMO)-based regimen (e.g. bendamustin); 4) monoclonal antibody (MAB)-based regimen (e.g. daratunumab). Direct medical costs of mandatory health insurance were analysed in 2017 Swiss Francs (CHF; third party payer perspective).

**Results:** Overall, we identified n=1054 prevalent MM patients (2012-2017) and n=378 incident MM patients (2015-2017; men: 47.1%; age group <=75 years: 48.7%). The number of prevalent patients per year increased over time (from n=314 in 2012 to n=645 in 2017).

PI-based regimens were the most frequent first line approach for incident patients (76.0%), followed by IMID-based (21.9%) and CHEMO-based regimens (2.1%). Only four patients were treated with MAB-drugs. For later lines, IMID-based regimens were most often used (2^nd^ line: 56.4%; 3^rd^ line: 2 of 3 patients), followed by PI-based regimens (43.6% and 1 of 3 patients, respectively). 161 of 1054 prevalent MM patients (15.3%) were treated with autologous hematopoietic stem cell transplantation (HSCT), 4 patients with allogeneic HSCT.

Average costs per patient per treatment line varied considerably (reliable data available from 2012 to 2014; mean duration of lines between 112 and 388 days): PI-based regimens: CHF 81’352; IMID-based: CHF 73’495; CHEMO-based: CHF 683. Mean daily costs under MM treatment stepwise increased from CHF 209 in 2012 to CHF 254 in 2017 (relative increase: 21.5%). Annual direct medical costs in Switzerland for seven novel MM drugs were extrapolated to be 60.1 Mio CHF in 2012 and 118.6 Mio CHF in 2017 (relative increase: 97.3%), corresponding to mean annual outpatient MM drug costs per patient of CHF 28’000 in 2017.

Annual death rates decreased systematically from 18.6% in 2012 to 15.5% in 2017 (p for trend: 0.03). No statistically significant difference in death rates emerged for 2017 compared with 2012 (risk ratio: 0.83; 95%-CI: 0.63 to 1.10; absolute risk reduction: 3.1%).

**Conclusions:** Current treatment patterns for MM patients in Switzerland show variation concerning applied drug regimens as well as costs. An increasing prevalent population of MM patients in combination with increasing costs per day under treatment lead to a substantial and growing budget impact for the Swiss social insurance system.

## Background

Multiple myeloma (MM) is the second most frequent hematologic malignancy in Switzerland. Incidence of MM is increasing since 1990 world-wide [1] and it presents a considerable burden to patients and the health care system. [2] Novel drugs (e.g. bortezomib, lenalidomide, carfilzomib) have considerably changed current treatment regimens for MM. Novel approaches have also improved prognosis of MM patients but are expensive. [2-4] Updated guidelines for Switzerland [2] and for Europe [3] advise clinicians regarding the translation of current evidence derived from clinical studies into treatment strategies in clinical practice to secure patient benefit.

Little is known, however, regarding the use of current MM treatment regimens in routine clinical practice, associated costs and mortality in the real-world setting. [5, 6] For Switzerland, data exist only from a pan-European study with very few Swiss patients included. [7, 8]

Health care claims data cover a large population and are increasingly used in health services research. They are usually reliable as they underly multiple check procedures and represent a well-established approach to collect and analyse real-world treatment data and costs. Such data allow quantifying the medical and economic burden of a disease to better understand current public health challenges and to optimise resource allocation. We therefore used health care claim-based data to describe the current real-world patterns and associated costs of MM treatment in Switzerland.

## Methods

### Study design

We performed a retrospective, observational study (cost-outcome description [9]). We used the claims database of consecutive cases of a major national Swiss health insurance company, which covers about 14% of the Swiss population (Helsana Versicherungen AG).

According to the Human Research Act in Switzerland, approval from a local Ethics committee was not required for studies using an anonymised database. The study was partly funded by Amgen Switzerland. The funding party had no influence on the concept of the study, data collection, analysis or interpretation of results.

### Study period and patients

Data were collected from the complete calendar years 2012 to 2017. To be included, cases had to be insured with Helsana health insurance for the complete calendar year (prevalence view). In addition, complete three preceding years without identification of MM were required to allow the inclusion as incident cases (incidence view). In case of death, cases were included even if their Helsana health insurance coverage was less than 12 months in that year.Thus, prevalent cases relate to the years 2012 to 2017, incident cases to the years 2015 to 2017 (Supplement: Figure S1). For analyses of treatment lines and costs, we relied on the incident population, because in this population we could unequivocally identify the first and subsequent treatment lines of each given case.

### Inclusion criteria

The used health insurance database holds no direct information concerning the diagnosis of their patients treated in the outpatient sector. Thus, we had to rely on an algorithm using primary and secondary inclusion criteria for MM. For patients with in-hospital treatment we used ICD10 diagnosis codes as primary identification (C90.0 Multiple myeloma).

For patients without hospitalisation and without the respective ICD code hospital discharge codes for identification of MM, we applied a filter algorithm for specific drugs to select patients suffering from MM with a very high probability (secondary inclusion criteria). Approved and reimbursed MM drugs in Switzerland (bortezomib, carfilzomib, ixazomib, lenaladomide, pomalidomide, elotuzumab, daratumumab) were identified via ATC-codes, as well as other drugs mostly used in the context of MM treatment (e.g. Bendamustin; Melphalan; Dexamathasone; Zoledronate). Of these identified cases, we excluded those with additional use of Rituximab (as an indicator for non-Hodgkin lymphoma), and with the use of iron chelators (e.g. desferoxamine, as an indicator for Myelodysplastic Syndrome).

### Information from claims database

Data were collected for the study population (per person per year) for demographic attributes (e.g. age, gender, death [yes/no]), health insurance attributes (e.g. managed care coverage [yes/no]), sequence of MM medications to identify treatment regimens, hematopoietic stem cell transplantation (HSCT: Swiss-DRG codes: A15C autologous; A04C allogeneic) and in-hospital stay due to MM.

Validated death data of the years 2012 and 2013 were taken from an earlier pilot study. This was necessary as a recoding for some variables in the insurance database took place during the observation period. Thus, prevalence figures used as denominators for calculation of death rates in 2012 and 2013 are not identical with prevalence figures reported in our study for other outcomes than death.

### Treatment patterns

We grouped MM treatments according to the major drug backbone into drug regimens (Table S1): The PI-based regimen for the proteasome inhibitor concept (e.g. bortezomib), the immunomodulatory (IMID)-based regimen (e.g. lenalidomide), the CHEMO-(chemo therapy-) based regimen (e.g. bendamustin) and the MAB-(monoclonal antibody)-based regimen (e.g. Elotuzumab) [10-12]. In addition, a hierarchy of regimens was defined (PI > IMID > CHEMO > MAB) to meaningfully group simultaneously prescribed MM drugs to MM regimens. For example, if bortezomib and lenalidomide were applied within one drug regimen, the regimen was defined as PI-based regimen; if lenalidomide and bendamustin were applied within one drug regimen, the regimen was defined as IMID-based regimen.

As a pragmatic compromise, we defined a new treatment line as a MM treatment regimen after a “watch and wait phase” of at least 180 days without prescription of a MM specific drug (i.e. PI-; IMID-; CHEMO-; MAB-drug). As no information was available in the claims database about the reasons for change of treatment (e.g. due to relapse or side effects) or treatment breaks (e.g. due to remission), we performed a validation study to better understand prescription patterns of MM drugs in our claims database. A visual inspection of diverse MM drug patterns of 16 example patients showed that a shorter period than 180 days (for example, 60 days) would erroneously detect a “new line” also in cases where clearly no new line was visible in the validation patterns. Current Swiss guidelines recommend repetition of drug regimens after 21 days to 6 weeks within a treatment line [2] and prescription can be given (and, thus, claims can exist) for several cycles together. For example, one prescription of 5 drug packages at a time of the same MM drug may hold for 5 therapy cycles within the same treatment line (5 × 28 days = 140 days). Consequently, the next prescription of the same drug within the same treatment line after 140 days would mimic a new line, if the limit would be set at, for example, 60 days and not at 180 days.

### Health economic analysis

Analysis was done from the perspective of a third party payer (Helsana Insurance AG). In Switzerland, in-hospital drug costs are in general included in the Swiss-DRG tariffs and not separately reimbursed by health insurers. In the outpatient sector, health insurers pay each drug prescription on a fee-for-service base.

Direct medical costs (e.g. total medication cost of MM medications listed in inclusion criteria) were defined from the database via number of reimbursed units (e.g. prescribed medication package) multiplied with current Swiss basic health insurance tariffs (OKP) per unit. Costs for in-hospital treatment in Switzerland are based on Swiss-DRG cost weights multiplied with a base rate (45% of in-hospital treatment are paid by the patients’ mandatory health insurance and included in our study; 55% by public authorities [cantons] and not included in our study). Costs are presented as 2017 Swiss Francs (CHF; official 2017 conversion rate to Euros: 0.85). [13] Out-of-pocket payments and deductibles of the patients were not accounted for. In addition, we did not apply a discount rate for costs due to the short observation period.

The extrapolation of costs to the Swiss population was performed with Helsana market shares per stratum as defined in the Swiss risk compensation scheme to adjust for differences across demographics between the Helsana population and the general Swiss population. [14] The applied attributes for defining the strata were age group, canton of residence and gender which result in 32 groups per canton of residence.

### Statistical analysis

For our descriptive analysis, we used means (standard deviation) or medians (interquartile range [IQR]) for continuous variables and proportions for categorical data. For inferential analysis, we applied parametric and non-parametric tests and p-values <0.05 were considered significant. R was used for data analysis: R version 3.5.0 (2018-04-23) - R Core Team (2017). R: A language and environment for statistical computing. R Foundation for Statistical Computing, Vienna, Austria.URL https://www.R-project.org/.

## Results

### Patient population

We identified n=1054 prevalent MM patients (2012-2017) with 3061 person-years under observation, and n=378 incident MM cases (2015-2017; study flow Figure 1). Patient characteristics and the number of patients per year of the two groups are depicted in Table 1. For prevalent patients, the number of patients per year increased over time (from n=314 in 2012 to n=645 in 2017), as well as patient age (age group >75 years: from 38.2% to 47.1%) and managed care coverage (from 33.8% to 49.6%). No clear pattern emerged for other variables. For example, the ratio of incident patients identified via ICD-code, hence with first diagnosis of MM in a hospital, varied between 70% and 82% from 2015 to 2017.

**Table 1:** Patient characteristics, mortality and indicators for co-morbidity of prevalent and incident MM patients. Analysis based on n=1054 patients from 2012-2017)

|  |  | n=1054 prevalent MM patients |  |  |  |  |  |  | n=378 incident patients |  |  |  |
| --- | --- | --- | --- | --- | --- | --- | --- | --- | --- | --- | --- | --- |
|  |  | Jahr 2012 | Jahr 2013 | Jahr 2014 | Jahr 2015 | Jahr 2016 | Jahr 2017 | p trend | Jahr 2015 | Jahr 2016 | Jahr 2017 | p trend |
| group size | n | 314 | 408 | 506 | 568 | 620 | 645 |  | 145 | 125 | 108 |  |
| age '<=75' | n (%) | 194 (61.8) | 243 (59.6) | 284 (56.1) | 312 (54.9) | 338 (54.5) | 341 (52.9) | < 0.01 | 69 (47.6) | 62 (49.6) | 53 (49.1) |  |
| age '>75' | n (%) | 120 (38.2) | 165 (40.4) | 222 (43.9) | 256 (45.1) | 282 (45.5) | 304 (47.1) | < 0.01 | 76 (52.4) | 63 (50.4) | 55 (50.9) |  |
| gender 'female' | n (%) | 159 (50.6) | 208 (51) | 255 (50.4) | 294 (51.8) | 311 (50.2) | 323 (50.1) | 0.57 | 77 (53.1) | 68 (54.4) | 55 (50.9) |  |
| gender 'male' | n (%) | 155 (49.4) | 200 (49) | 251 (49.6) | 274 (48.2) | 309 (49.8) | 322 (49.9) | 0.57 | 68 (46.9) | 57 (45.6) | 53 (49.1) |  |
| managed care coverage 'yes' | n (%) | 106 (33.8) | 148 (36.3) | 190 (37.5) | 240 (42.3) | 288 (46.5) | 320 (49.6) | < 0.01 | 66 (45.5) | 58 (46.4) | 59 (54.6) |  |
| deceased 'yes' | n (%) | 68/366 (18.6)* | 73/358 (20.4)* | 89 (17.6) | 93 (16.4) | 99 (16.0) | 100 (15.5) | 0.03 | 20 (13.8) | 16 (12.8) | 12 (11.1) |  |
| identification via C90 'yes' | n (%) | 191 (60.8) | 290 (71.1) | 422 (83.4) | 468 (82.4) | 509 (82.1) | 505 (78.3) | 0.11 | 114 (78.6) | 102 (81.6) | 76 (70.4) |  |
| nr. of different ATC | mean (median) | 22.9 (22) | 22.5 (22) | 21.9 (21) | 21.7 (21) | 22.1 (21) | 21.1 (20) | 0.02 | 26.2 (26) | 27.2 (27) | 25.6 (25) |  |
| nr. of PCG | mean (median) | 4.8 (5) | 4.7 (5) | 4.5 (5) | 4.6 (4.5) | 4.5 (4) | 4.4 (4) | 0.01 | 5.3 (5) | 5.4 (5) | 5.4 (5) |  |
| PI-based regimen | n (%) |  |  |  |  |  |  |  | 100/119 (84.0) | 73/90 (81.1) | 49/83 (59.0) | 0.27 |
| IMiD-based regimen | n (%) |  |  |  |  |  |  |  | 16/119 (13.4) | 15/90 (16.7) | 33/83 (39.8) | 0.26 |
| CHEMO-based regimen | n (%) |  |  |  |  |  |  |  | 3/119 (2.6) | 02/90 (2.2) | 01/83 (1.2) | 0.15 |

\*Death rates: Validated death data of the years 2012 and 2013 were taken from an earlier pilot study. This was necessary as a recoding for some variables in the insurance database took place during the observation period. Thus, prevalence figures used as denominators for calculation of death rates in 2012 and 2013 are not identical with prevalence figures reported in our study for other outcomes than death.
ATC: WHO Anatomical therapeutic chemical (ATC) classification system for drugs, an indicator for co-morbidity
PCG: pharmacy cost groups; an indicator for co-morbidity

**Figure 1:**
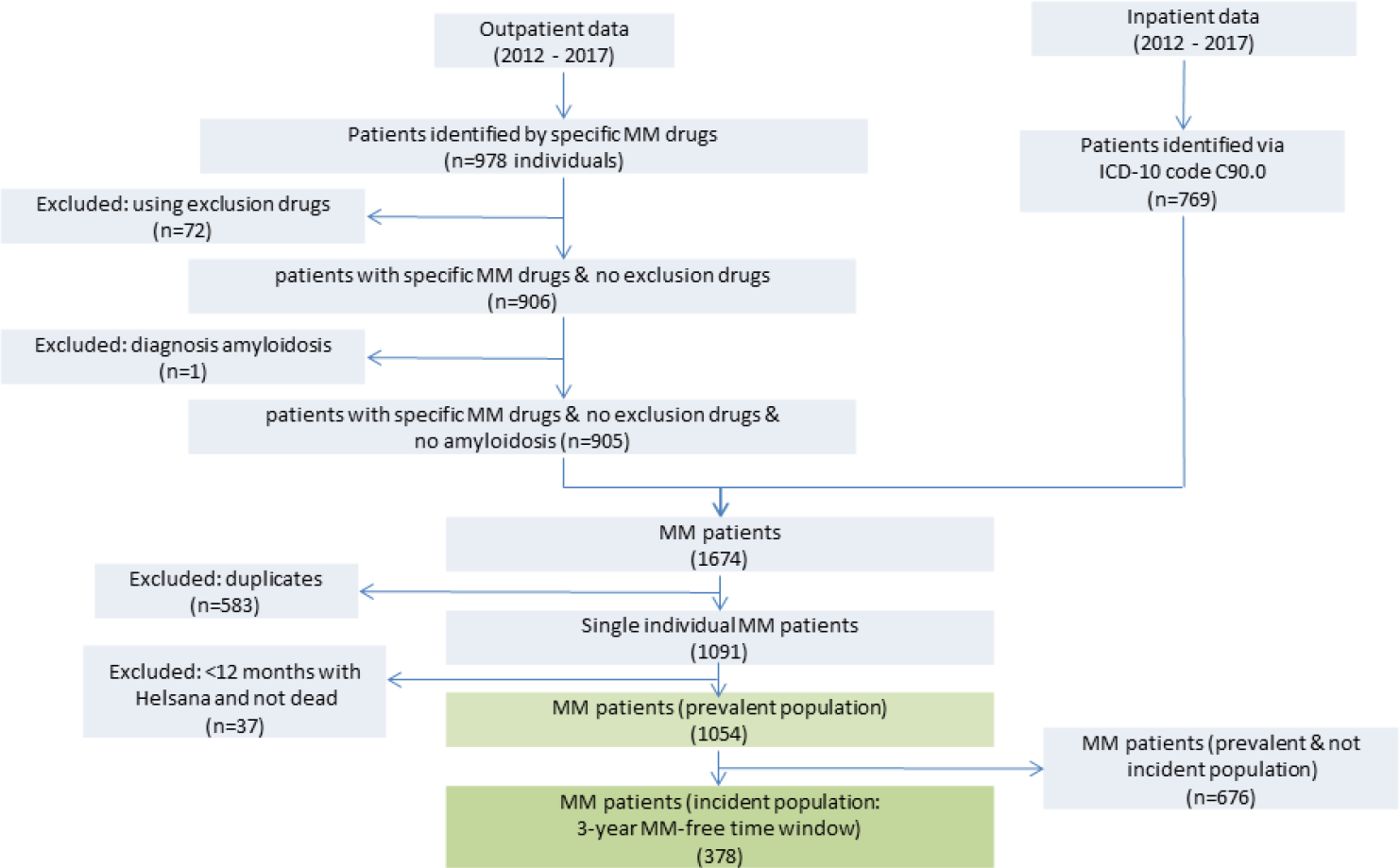
Study flow.

When we extrapolated the figures derived from the Helsana sample to the Swiss population, figures for 2017 did correspond to 61.0 prevalent cases per 100’000 inhabitants (95%-CI: 58.3 to 63.8) and 10.5 incident cases / 100’000 (95%-CI: 8.6 to 12.5).

### Drug treatment patterns

Of 378 incident patients, 292 (77.2) provided outpatient drug data with a mean follow-up of 2.1 years between 2015 and 2017 to assess drug treatment in further detail. For 1^st^-line therapy, PI-based regimens were the most frequent approach (76.0%), followed by IMID-based (21.9%) and CHEMO-based regimens (2.1%; Figure 2). Only four patients were treated with MAB-drugs, three times as part of PI-based regimens and once as part of an IMID-based regimen.

**Figure 2:**
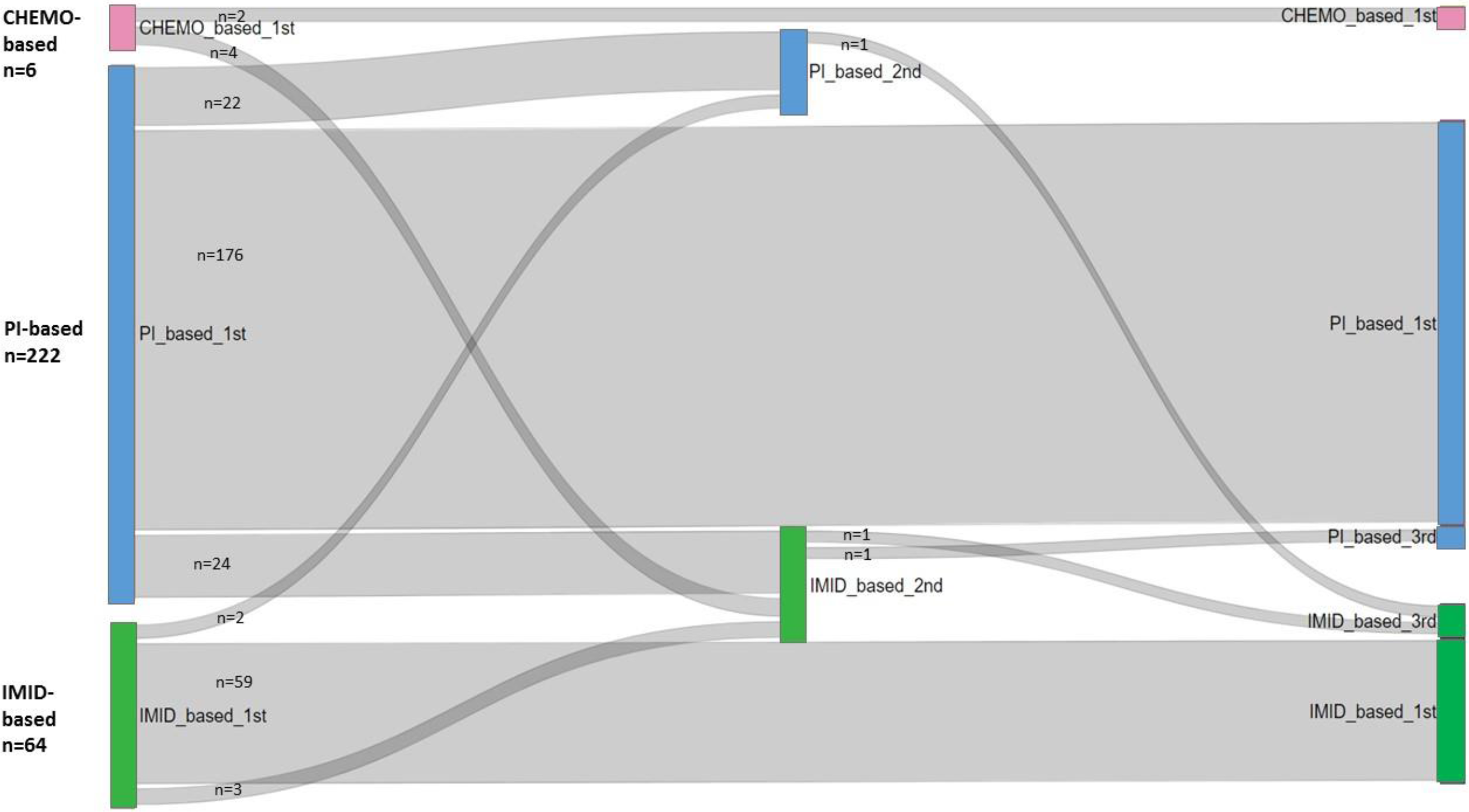
Treatment patterns. Sequence of treatment lines according to 1st-line drug regimen groups (PI-based: mean duration of 1^st^ line: 218 days; IMID-based: 214 days; CHEMO-based: 64 days). Analysis based on 292 of 378 incident MM patients from 2015 to 2017.

Age distribution across 1^st^-line therapies showed a decrease of the younger age population from PI-based (age group <=75 years: 59%), over IMID-based (48%) to CHEMO-based (17%) regimens (Table 2).

**Table 2:** First line treatment patterns, patient variables, mortality and HSCT for MM patients. Analysis based on 292 of 378 incident MM patients with outpatient drug data from 2015-2017.

| **2015-2017: Regimens** |  | **PI-based** |  | **IMiD-based** |  | **CHEMO-based** |  | **TOTAL** |  |
| --- | --- | --- | --- | --- | --- | --- | --- | --- | --- |
| group size | N | **222** | **76%** | **64** | **22%** | **6** | **2%** | **292** | **100%** |
| group size | n | 222 | 100% | 64 | 100% | 6 | 100% |  |  |
| age '<=75' | n | 131 | 59% | 31 | 48% | 1 | 17% | 163 | 56% |
| age '>75' | n | 91 | 41% | 33 | 52% | 5 | 83% | 129 | 44% |
| gender 'female' | n | 109 | 49% | 39 | 61% | 4 | 67% | 152 | 52% |
| gender 'male' | n | 113 | 51% | 25 | 39% | 2 | 33% | 140 | 48% |
| managed care coverage 'yes' | n | 110 | 50% | 33 | 52% | 2 | 33% | 145 | 50% |
| deceased 'yes' | n | 13 | 6% | 4 | 6% | 1 | 17% | 18 | 6% |
| HSCT any time 'yes' | n | 54 | 24% | 4 | 6% | 0 | 0% | 58 | 20% |
| HSCT in incidence year 'yes' | n | 36 | 16% | 4 | 6% | 0 | 0% | 40 | 14% |

HSCT: Hematopoietic stem cell transplantation

Mean duration of 1^st^-line treatment in 2015 and 2016 was between 210 and 298 days for PI-based regimens, between 197 and 327 days for IMID-based regimens and between 30 and 61 days for CHEMO-based regimens (Table S2).

Mean time to next treatment in 2015 and 2016 (TTNT; i.e. interval from last day of 1^st^-line to first day of 2^nd^-line treatment) was between 283 and 351 days for PI-based regimens, and between 216 and 389 days for IMID-based regimens. Only 5 patients provided data for CHEMO-based regimens (TTNT: 207 to 390 days). Duration of treatment and TTNT in 2017 were generally shorter, most probably due to the end of the study time window by December 2017.

During the time window between 2015 and 2017 (mean follow-up: 2.1 years), 237 (81.2%) patients remained in their 1^st^-line treatment. For 55 of 292 patients (18.8%) a 2^nd^-line treatment was started. The most frequent 2^nd^-line regimen was IMID-based (56.4%), followed by PI-based (43.6%) and no CHEMO-based 2^nd^ line. Three of 292 patients (1%) were also treated with a 3^rd^-line regimen during our time window (2 of 3 with IMID-based; 1 of 3 with PI-based; no CHEMO-based regimen).

### Hematopoietic stem cell transplantation

From 2012 to 2015, 161 of 1054 MM patients (15.3%) were treated with autologous HSCT, four patients were treated with allogeneic HSCT (Table S3). All patients with HSCT were younger than 75 years with a balanced gender distribution as in the total prevalence population of 1054 MM patients.

### Deceased MM patients

The proportion of deceased patients per year showed a systematically decreasing trend (p for trend: 0.03) during the observation time from 2012 to 2017 (Table 1): 68 of 366 patients (18.6%) died in 2012, 73 of 358 patients (20.4%) in 2013, 89 of 506 patients (17.6%) in 2014, 93 of 568 (16.4%) in 2015, 99 of 620 (16.0%) in 2016 and 100 of 645 (15.5%) in 2017. No statistically significant difference in death rates emerged for 2017 compared with 2012 (risk ratio: 0.83; 95%-CI: 0.63 to 1.10; absolute risk reduction: 3.1%; Figure 3).

**Figure 3:**
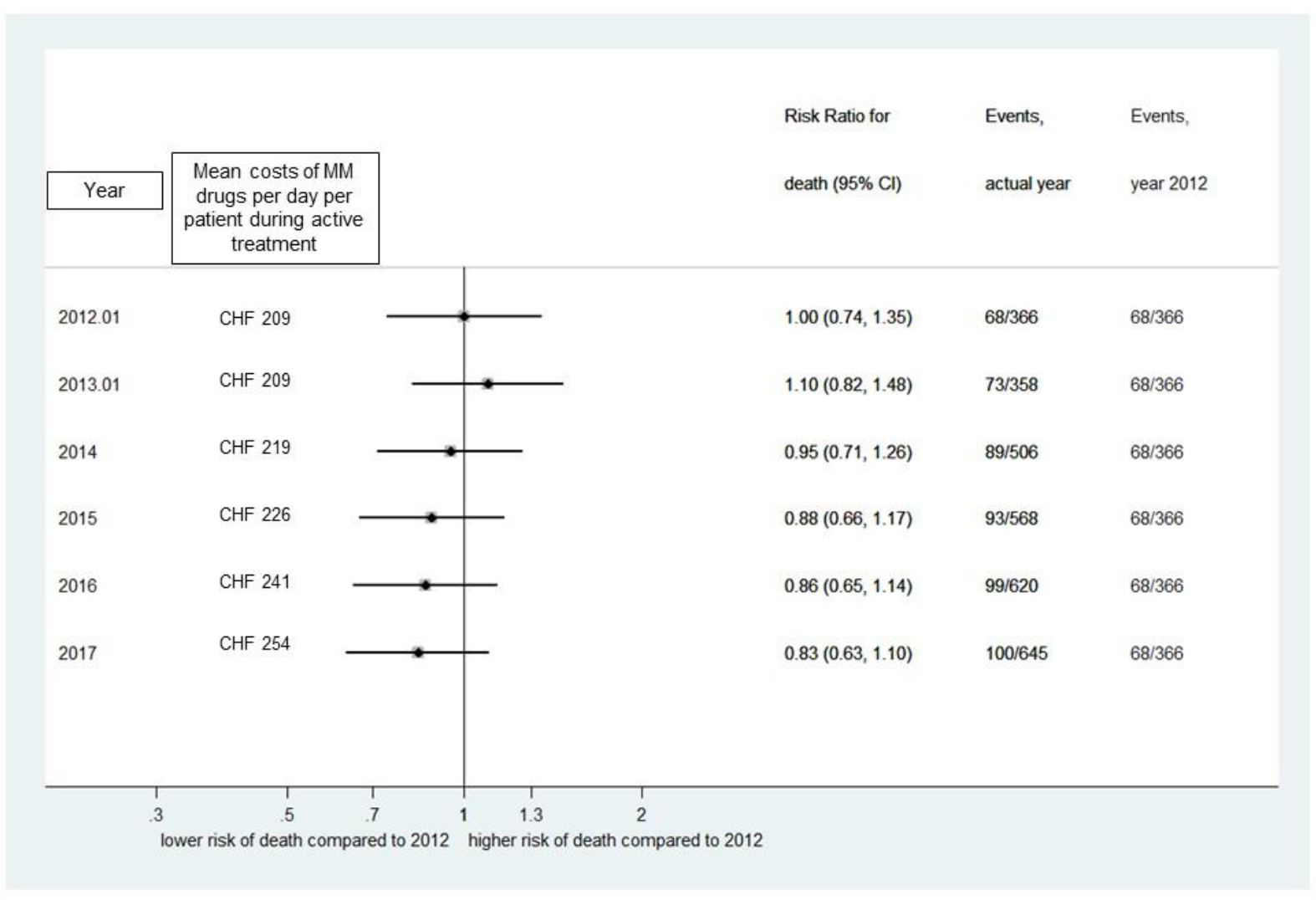
Risk ratio for death and daily treatment costs under MM treatment compared to 2012. Analysis based on 3061 observed patient years of prevalent MM patients from 2012 to 2017. All risk ratios are calculated for reference year 2012.

### Treatment costs for MM

Direct medical costs for MM specific drugs per patient per treatment regimen varied considerably between regimens, as well as between years (Table 3). Valid cost data for complete treatment lines were available from 2012 to 2014. Cost data of treatment lines from 2015 to 2017 are not fully reliable due probably many incomplete treatment lines at the end of observation period. From 2012 to 2014, mean costs for PI-based treatment lines varied between CHF 74’315 and CHF 94’914 and for IMID-based lines between CHF 59’227 and CHF 85’169. Mean costs for CHEMO-based regimens were considerably less costly (CHF 285 to CHF 1235). Mean costs per patient per day under a MM drug regimen were available from 2012 to 2017 (formula: average costs per regimen / average number of days under a regimen; watch and wait periods without MM drugs are not included herein). Mean daily costs stepwise increased from CHF 209 in 2012 to CHF 254 in 2017 (relative increase: 21.5%).

**Table 3:** Outpatient MM drug costs. Mean costs per MM drug regimen: Regimens are from any treatment line. The first day of drug regimen defines costing year. Analysis is based on 3061 observed patient years of prevalent MM patients from 2012 to 2017. From 2015 onwards, no valid cost data are available for complete treatment lines (light grey figures), as probably many treatment lines are not fully covered until end of observation period. Mean costs of MM drugs per day per patient (formula: average costs per regimen / average number of days under a regimen; watch and wait periods without MM drugs are not included herein). Valid cost data per day are available from 2012 to 2017.

|  | 2012 |  | 2013 |  | 2014 |  | 2015 |  | 2016 |  | 2017 |  |
| --- | --- | --- | --- | --- | --- | --- | --- | --- | --- | --- | --- | --- |
|  | CHF |  | CHF |  | CHF |  | CHF |  | CHF |  | CHF |  |
| Mean costs per MM drug regimen | mean | 95% - CI | mean | 95% - CI | mean | 95% - CI | mean |  | mean |  | mean |  |
| all regimens | **73764** | 137 to 397007 | **76368** | 85 to 374417 | **71060** | 187 to 313594 | **60297** | 274 to 238801 | **66103** | 1397 to 225936 | **34085** | 1475 to 98029 |
| PI_based | 74827 | 1411 to 342232 | 94914 | 3666 to 411318 | 74315 | 1741 to 324722 | 56673 | 1811 to 247540 | 68558 | 3686 to 225025 | 35508 | 1837 to 100327 |
| IMID_based | 85169 | 7440 to 447372 | 59227 | 6020 to 246112 | 76088 | 7128 to 302585 | 82800 | 6665 to 235718 | 65785 | 7001 to 236983 | 34384 | 5342 to 92911 |
| CHEMO_based | 528 | 52 to 3441 | 285 | 45 to 1289 | 1235 | 25 to 7382 | 270 | 49 to 588 | 400 | 50 to 1131 | 1049 | 57 to 3625 |
| Mean costs of MM drugs per day per patient |  |  |  |  |  |  |  |  |  |  |  |  |
| all regimens | **209** |  | **209** |  | **219** |  | **226** |  | **241** |  | **254** |  |
| PI_based | 202 |  | 213 |  | 214 |  | 217 |  | 229 |  | 235 |  |
| IMID_based | 243 |  | 212 |  | 239 |  | 257 |  | 273 |  | 282 |  |
| CHEMO_based | 3 |  | 3 |  | 21 |  | 3 |  | 16 |  | 13 |  |

Inpatient costs of patients with autologous HSCT in the year of transplantation show some variability from 2012 to 2017 (range: CHF 24’597 - CHF 33’090; mean inpatient all diagnosis costs per patient per year; Table S3).

We extrapolated annual outpatient MM drug costs to the general population of Switzerland (Table 4). Annual direct medical costs in Switzerland for seven novel approved and reimbursed MM drugs (bortezomib [Velcade®], lenalidomide [Revlimid®], pomalidomide [Imnovid®], carfilzomib [Cyprolis ®], elotuzumab [Empliciti ®], daratumumab [Darzalex ®], ixazomib [Ninlaro ®]) were extrapolated to be 60.1 Mio CHF in 2012 and 118.6 Mio CHF in 2017 (relative increase: 97.3%). In 2017, the relative share of costs in Switzerland among the four most frequently used novel agents was 55.3% for lenalidomide, 18.8% for bortezomib, 11.3% for pomalidomide and 7.1% for carfilzomib.

**Table 4:** Extrapolated annual outpatient MM drug costs of MM patients in Switzerland from 2012 to 2017. Extrapolation to Switzerland is based on 1054 prevalent Helsana MM patients from 2012 to 2017. Costs include only outpatient MM drugs as reimbursed by the Swiss Federal Law on Basic Health Insurance (BHI). Costs of inpatient treatment, of other drugs or of additional services are not included. MM drug costs are from any treatment line. Prescription date of each MM drug defines costing year. MM drugs are reported from 2012 onwards or from the first year of reimbursement in Switzerland. Costs (CHF; per patient per year): calculated for each calendar year, irrespective of active MM treatment; thus, watch and wait periods without MM drugs are included herein.

|  | 2012 |  | 2013 |  | 2014 |  | 2015 |  | 2016 |  | 2017 |  |
| --- | --- | --- | --- | --- | --- | --- | --- | --- | --- | --- | --- | --- |
|  | thousand CHF |  | thousand CHF |  | thousand CHF |  | thousand CHF |  | thousand CHF |  | thousand CHF |  |
|  | mean | 95% - CI | mean | 95% - CI | mean | 95% - CI | mean | 95% - CI | mean | 95% - CI | mean | 95% - CI |
| Lenaladomid (REVLIMID®) | 41471 | 32606 to 50337 | 37017 | 30017 to 44018 | 34811 | 28252 to 41370 | 41751 | 33756 to 49746 | 59687 | 50184 to 69189 | 65570 | 55746 to 75393 |
| Bortezomib (VELCADE®) | 18630 | 15156 to 22105 | 21382 | 17709 to 25056 | 24627 | 20308 to 28946 | 26963 | 22714 to 31211 | 24397 | 19028 to 29766 | 22292 | 17426 to 27158 |
| Pomalidomid (IMNOVID®) |  |  |  |  | 3945 | 1751 to 6139 | 12581 | 7103 to 18060 | 11579 | 5825 to 17333 | 13369 | 6908 to 19830 |
| Carfilzomib (KYPROLIS®) |  |  |  |  |  |  | 123 | 30 to 287 | 7370 | 4360 to 10379 | 8372 | 4752 to 11992 |
| Daratumumab (DARZALEX®) |  |  |  |  |  |  |  |  |  |  | 6826 | 3355 to 10298 |
| Ixazomib (NINLARO®) |  |  |  |  |  |  |  |  |  |  | 1787 | 294 to 3279 |
| Elotuzumab (EMPLICITI®) |  |  |  |  |  |  |  |  | 31 | 7 to 94 | 402 | 56 to 955 |
| Total | 60101 |  | 58399 |  | 63383 |  | 81418 |  | 103064 |  | 118618 |  |
| Estimated prevalence Switzerland (n) | 1810 |  | 2360 |  | 3030 |  | 3410 |  | 3910 |  | 4200 |  |
| Costs (CHF; per patient per year) | 33205 |  | 24745 |  | 20918 |  | 23876 |  | 26359 |  | 28242 |  |
| Costs total BHI (CHF; per pat per yr) | 68374 |  | 56941 |  | 55161 |  | 54820 |  | 58101 |  | 57006 |  |
| % costs MM-drugs of costs total BHI | 0.49 |  | 0.43 |  | 0.38 |  | 0.44 |  | 0.45 |  | 0.50 |  |

BHI: (Swiss) Basic Health Insurance; CHF: Swiss Francs; MM: multiple myeloma; pppy: per patient per year; Costs total OKP (CHF; pppy): include all-diagnoses inpatient and outpatient costs of myeloma patients, as reimbursed by health insurers based on the Swiss Federal Law on Basic Health Insurance (KVG/LAMal);

This corresponds to mean annual outpatient MM drug costs per patient of CHF 28’242 in 2017 (watch and wait periods without MM drugs are included herein; Table 4). In the period 2012 to 2017, annual costs of outpatient MM drugs (mean: CHF 26’224) accounted for about 45% of total annual costs of all-diagnoses inpatient and outpatient services of myeloma patients (mean: CHF 58’401), as reimbursed by health insurers based on the Swiss Federal Law on Basic Health Insurance (KVG/LAMal).

## Discussion

In our observational study of a claims database we analysed real-world treatment data of unselected MM patients in Switzerland in the era of novel MM drugs. PI-based regimens were the by far most frequent 1^st^-line therapy, followed by IMID-based and CHEMO-based regimens. Duration of 1^st^-line treatment in 2015 and 2016 was between 210 and 298 days for PI-based regimens, between 197 and 327 days for IMID-based regimens and between 30 and 61 days for CHEMO-based regimens. Time to next treatment (TTNT; i.e. interval from last day of 1^st^-line to first day of 2^nd^-line treatment) in 2015 and 2016 was between 283 and 351 days for PI-based regimens, and between 216 and 389 days for IMID-based regimens. For 2^nd^ and 3^rd^-line regimens IMID-based regimens are more frequent compared to PI-based regimens. 15.3 % of patients were treated with autologous ASCT.

Outpatient costs for MM drugs were highest for PI-based regimens (average costs for treatment lines started between 2012 and 2014: CHF 81’352), followed by IMID-based regimens (CHF 73’495). Costs for CHEMO-based regimens were much lower (CHF: 683), but patients treated with such regimens represent a specific group (only six patients in our data set with five of them in the age group >75 years).

Extrapolated costs of novel MM drugs to the Swiss national level increased by 97% from 2012 to 2017. Annual mortality rates of MM patients in the prevalent population of this sample of a Swiss health insurance from 2012 to 2017 decreased slightly from 18.6% to 15.5% (risk ratio: 0.83; 95%-CI: 0.63 to 1.10).

### Strengths and limitations

We applied real-world data of a major Swiss health insurance that provided treatment information about unselected MM patients from all parts of the country. The database has been used in several health services research studies in different clinical domains. [15-17]

Our study has, however, several limitations. First, we were not able to use clinical charts of single patients to verify diagnosis, provide information about staging or analyse MM drug treatment and new treatment lines directly from chart reviews in detail (e.g. to study separation of induction and maintenance therapy or reasons for ending treatment). Thus, we were only able to include MM patients who had ever received MM drug treatment or have been treated for MM in a hospital. However, our filter criteria were developed in cooperation with clinical experts and the chosen filter variables also used ICD-coded diagnoses of inpatient treatments. For definition of a new treatment line we used a pause of drug prescription of 180 days as a pragmatic approach in the light of Swiss guidance for MM treatment [2] and the reported real-world drug prescription modes by our clinical experts. Second, the chosen definition of three calendar years free of a MM drug prescription or an in-hospital stay without MM diagnosis for incident cases may still have been too short. Thus, we cannot exclude a misclassification of some prevalent cases as incident cases but this may be a rare event. Third, we do not recommend using the extrapolated MM prevalence and incidence figures for epidemiological questions. Our extrapolated estimates are likely to be too high as compared with published data that were properly adjusted for European standard populations. Fourth, we did not have direct information about co-morbidities of patients. However, using the validated ATC-codes for medically treated co-morbidities, [15] we have evidence, that the co-morbidity profile of outpatients remained basically similar over the observation period. Fifth, some results may not be reliable due to a short observation time window for patients who entered the study late. For example, for the analysis of the sequence of treatment lines, study population with suitable data is decreasing rapidly from line to line. Furthermore, TTNT may be underestimated for patients who entered the sample in 2017 due to the relatively short time window until end of observation period. Finally, the Helsana population is somewhat older than the Swiss general population and not all parts of Switzerland were equally represented in our study. However, extrapolating the costs to the country level we accounted for age differences, canton of residence and gender.

### Recent publications with real-world data of MM patients

We compared our findings with recently published real-world treatment data of MM patients. An observational study from 6 countries [7, 8] included also data from one Swiss hemato-oncological centre: These cross-country findings are, however, difficult to compare with our results, as the European authors performed chart reviews of patients seen during consultation and could, thus, provide detailed clinical information but no figures about mortality rates and costs.

In the multi-country study [7], the rate of patients receiving a 2^nd^-line (61%) and a 3^rd^-line therapy (38%) is higher compared with the findings in our incident population (2^nd^-line: 19%; 3^rd^-line therapy: 1%). This may be partly explained with the longer time since diagnosis (median: 33 months) in the multi-national sample. [7] In addition, stem cell transplantation is less frequent in our sample (15% vs. 31%). Treatment patterns are similar with the most frequent 1^st^-line drug bortezomib (EU: 48%; PI-based regimen in our analysis: 76%). Lenalidomid is the most frequent drug for 2^nd^-line (EU: 59%; IMID-based regimen in our analysis 56%) and 3^rd^-line therapy (EU: 63%; our findings: 67%. [7] A real world study from the US with data from 2006 to 2014 found a similar pattern. [6] Treatment free intervals between lines (TTNT) in the EU study are between 10 months (median; after 1^st^ line) and 5 months (after 2^nd^ line). [8] We found a mean TTNT after 1^st^-line between 7 and 13 months, for IMID- and PI-based regimens in 2015 and 2016).

A real-world study from the Netherlands assessed treatment costs in MM patients. [5] Even though this study assessed costs between 2001 and 2009, acquisition costs for novel MM agents accounted for about one third of total monthly inpatient and outpatient costs in the Netherlands at that time. We did not calculate total health care costs per patients of our sample, as we were mainly interested in diagnosis specific costs of MM treatment. However, it is likely that the increase in costs in our study for novel MM drugs in Switzerland from 2012 to 2017 accounts for more than one third of treatment costs in our MM patients.

HSCT was performed in 15.3% of prevalent patients in our study. This is a similar fraction as in other real-world populations from the US [6] (data from 2006 to 2014: 16.2%) or from the Netherlands [5] (data from 2001 to 2009: 20%). The European multi-country study [7] showed a higher fraction of HSCT (data from 2014: 31%), which may be due to an over-representation of patients from University oncology centers.

### Implications for clinicians and public health decision makers

Our study provides evidence, that initial drug treatment regimens show little variability in accordance with current guidance. [2] However, variability of treatment patterns is increasing for 2^nd^- and 3^rd^-line therapy in our real-world data. Unexpected toxicities or patient preferences may contribute to such modifications.

Our study shows a significant increase in daily drug costs for MM patients over a six year period, on the patient level as well as on the Swiss country level. For example, the introduction of new agents, such as pomalidomide or monoclonal antibodies, was not associated with a decrease in costs for other myeloma drugs. The doubling of costs for novel MM drugs on the national level (relative increase by 97% from 2012 to 2017), is probably mainly due a massive increase of the prevalent MM population (as indicated by a relative increase of MM patients from 2012 to 2017 in our data set of 105%). This may represent the better prognosis of MM patients and may not be due to a rise in MM incidence, as indicated in our data set. To a lower extend, also increasing MM drug costs per patient per day have contributed to a higher estimated spending on the national level. The relative increase of 15% of MM drug costs per day (CHF 209 in 2012; CHF 241 in 2016) was similar to the relative increase of 16% in overall health care expenditures in Switzerland during the period 2012 to 2016 [no official data for 2017 available, yet]). [18]

Surprisingly, the application of novel MM drugs is not (yet?) associated with a significant decrease in mortality among MM patients from 2012 to 2017. However, we found a significant trend towards reduced mortality. Admittedly, we have only little clinical information about the MM patients in our sample, but no reason to believe, that these MM patients are, beyond the slightly increased age, significantly different compared to a typical MM population in Switzerland. Our data are in line with the US SEER registry, where mortality from myeloma in the US population has been slightly decreasing over the years from 2010 to 2015 in the era of novel MM drugs. [19] Data from other hematologic diseases have shown decreased mortality rates also on the population level after introduction of novel drugs. As an example may serve the effect of imatinib on survival for patients with chronic myeloid leukemia, where death rates in the US in the SEER registry dropped from 0.6 per 100’000 in 2000, the year of introduction of imatinib, to 0.3 in 2007, where they remained since despite introduction of newer tyrosine kinase inhibitors. [19]

Our data on novel MM drugs in Switzerland call for an introduction of “Coverage with Evidence Development” (CED) reimbursement. [20] This form of reimbursement provides continued access to novel MM drugs in Switzerland and might trigger the establishment of registries and research. Thus, a framework will be established to systematically assess and better understand the impact on patients’ outcome and costs in real-world settings. Monitoring such data on the population level is of outmost importance, to assess if novel MM drugs are associated with increased value for health care. [21]

## Conclusions

Current treatment patterns for MM patients in Switzerland show variation concerning applied drug regimens as well as costs. An increasing prevalent population of MM patients in combination with increasing costs per day under treatment lead to a substantial and growing budget impact for the Swiss social insurance system.

## Data Availability

The health insurance claims data as used for our analysis are stored at Helsana Versicherungen AG, Switzerland. Any questions concerning these data should be directed to EB.

## Glossary

|  |  |
| --- | --- |
| MM drug | One defined pharmacological substance for MM treatment (e.g. BOR, LEN, BENDA) |
| Treatment regimen | Combination of several MM drugs (e.g. PI-based regimen, IMID-based regimen; CHEMO-based regimen; MAB-based regimen) |
| Treatment line [22] | Cycle of a single MM agent or a regimen of several MM drugs or regimens (e.g. given as 1<sup>st</sup>-line treatment; 2<sup>nd</sup>-line treatment; 3<sup>rd</sup>-line treatment) |
| Treatment pattern | Composition and sequence of MM treatments over time |

## List of abbreviations

BENDA: bendamustin
BOR: bortezomib (e.g. Velcade ®)
CHEMO: chemotherapy-based regimen
DARA: daratunumab (e.g. Darzalex ®)
DEXA: dexamethason
DOXO: doxorubicin
ELO: elotuzumab (e.g. Empliciti ®)
HSCT: hematopoietic stem cell transplantation
IMID: immunomodulatory-based regimen
IXA: ixacomib (e.g. Ninlaro ®)
LEN: lenalidomide (e.g. Revlimid ®)
MEL: melphalan
PI: proteasome inhibitor-based regimen
POM: pomalidomide (e.g. Imnovid ®)
PRED: prednisolon
THAL: thalidomide
VIN: vincristine

## Declarations

### Ethics approval and consent to participate

Not applicable. According to the Human Research Act in Switzerland, approval from a local Ethics committee was not required for studies using an anonymised database.

### Authors’ Contributions

KE is the guarantor and drafted the manuscript. All authors contributed to the development of the selection criteria and data extraction criteria. RR, OR and EB managed the claims database and extracted data. RR provided statistical expertise and performed the analysis. All authors contributed to interpretation of results. All authors read, provided feedback and approved the final manuscript.

## Acknowledgements

We established a scientific Advisory Board that comprised three experienced Swiss hemato-oncologists. The authors are grateful for their methodological support (in alphabetical order):

- From 2015 to 2018: Christoph Driessen, MD PhD, Department of Medical Oncology and Hematology, Cantonal Hospital St. Gallen, Switzerland
- Alois Gratwohl, MD PhD, Medical Faculty, University of Basle, Basle, Switzerland
- From 2015 to 2017: Thilo Zander, MD, Department of Medical Oncology, Cantonal Hospital Lucerne, Switzerland

Depending on the data available at that time point, the methodological comments of the Advisory Board were used for improving the present working paper. Additional methodological limitations and future recommendations, as indicated by the Advisory Board, are part of the “Strengths and Limitations” section of this working paper.

## Supplement

**Figure S1:**
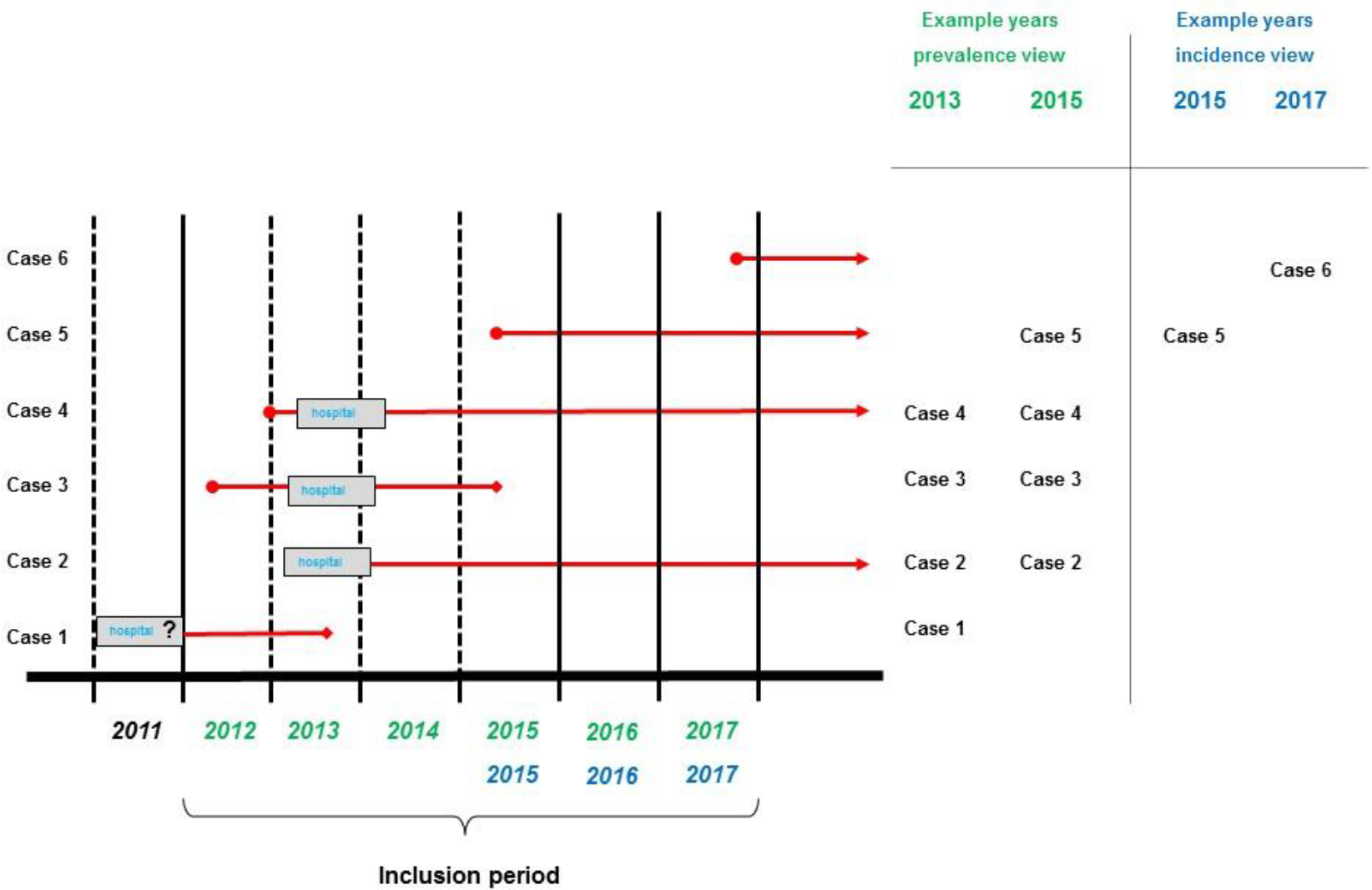
Inclusion procedure for prevalent and incident MM patients. Red lines are showing the trajectory of disease. Included persons had to be insured with Helsana for the whole calendar year.

**Table S1:** Applied grouping of MM drugs according to drug concepts into three MM drug regimens.

| drug concept | regimen | category | example drug combination (within regimen) |
| --- | --- | --- | --- |
| Proteasoma Inhibitor | PI-based | BOR | BOR +/- DEXA |
|  |  | BOR + IMID | BOR - THAL |
|  |  |  | BOR - LEN |
|  |  | BOR + CHEMO | BOR - MEL |
|  |  |  | BOR - Cyclophosphamid |
|  |  |  | BOR - liposomales DOXO (Caelyx®) |
|  |  |  | BOR - DOXO |
|  |  |  | BOR - BENDA |
|  |  | CAR + IMID | CAR - LEN |
|  |  | IXA |  |
| Imide | IMID-based | THAL | THAL - DEXA |
|  |  | LEN | LEN - DEXA |
|  |  | POM | POM - DEXA |
|  |  | LEN + CHEMO | LEN - MEL |
|  |  |  | LEN - BENDA |
|  |  |  | LEN - Cyclophosphamid |
|  |  | POM + CHEMO | POM - Cyclophosphamid |
|  |  | THAL + CHEMO | THAL - MEL |
|  |  |  | THAL - BENDA |
| Chemotherapy | CHEMO-based | BENDA | BENDA - DEXA |
|  |  | MEL | MEL - PRED |
|  |  | VIN | VIN |
| Monoclonal antibodies | MAB | Daratunumab | DARA - LEN- DEXA |
|  |  | Elotuzumab | ELO - POM - DEXA |

Abbreviations (in alphabetical order): BENDA-Bendamustin; BOR-Bortezomib; DARA-Daratunumab; DEXA-Dexamethason; DOXO-Doxorubicin; ELO-Elotuzumab; IXA-Ixazomib; LEN-Lenalidomid; MEL-Melphalan; POM-Pomalidomide; PRED-Prednisolon; THAL-Thalidomid; VIN – Vincristin

**Table S2:** Duration of first line treatment and time to next treatment (TTNT) in days. Analysis based on 292 of 378 incident MM patients from 2015 to 2017. Duration of treatment: days between first and last medication of first line treatment. TTNT: interval between the last day of 1st treatment line and the first days of 2nd treatment line (or last day of observation period).

|  |  | Jahr 2015 | Jahr 2016 | Jahr 2017 |
| --- | --- | --- | --- | --- |
| **total** |  |  |  |  |
| group size | n | 119 | 90 | 83 |
| duration of first line treatment | mean (days) | 221.8 | 275.2 | 135.5 |
| time between treatments 1 and 2 | mean (days) | 355.7 | 259.3 | 189 |
| time between treatments 1 and 2 or end of observation | mean (days) | 359.9 | 184.7 | 41.3 |
|  |  | Jahr 2015 | Jahr 2016 | Jahr 2017 |
| **PI-based** |  |  |  |  |
| group size | n | 100 | 73 | 49 |
| duration of first line treatment |  |  |  |  |
| total | mean | 209.8 | 298 | 146.6 |
| age <= 75 | mean | 189.5 | 328.2 | 137.9 |
| age > 75 | mean | 236.6 | 249.6 | 159.3 |
| HSCT yes a) | mean | 145.2 | 309.9 | 161.2 |
| HSCT no a) | mean | 232.4 | 294.4 | 142.4 |
| HSCT yes b) | mean | 133.8 | 287.8 | 161.2 |
| HSCT no b) | mean | 225.3 | 299.3 | 142.4 |
| time between treatments 1 and 2 | mean | 351.2 | 283.4 | 189 |
| time between treatments 1 and 2 or end of observation | mean | 358.2 | 185.5 | 43.2 |
| **IMiD-based** |  |  |  |  |
| group size | n | 16 | 15 | 33 |
| duration of first line treatment |  |  |  |  |
| total | mean | 326.9 | 196.8 | 120.1 |
| age <= 75 | mean | 328.3 | 187.8 | 143.5 |
| age > 75 | mean | 325 | 210.3 | 105 |
| HSCT yes a) | mean | - | 109.3 | 133 |
| HSCT no a) | mean | 326.9 | 218.7 | 119.7 |
| HSCT yes b) | mean | - | 109.3 | 133 |
| HSCT no b) | mean | 326.9 | 218.7 | 119.7 |
| time between treatments 1 and 2 | mean | 389 | 215.5 | - |
| time between treatments 1 and 2 or end of observation | mean | 337.9 | 177.5 | 38.8 |
| **CHEMO-based** |  |  |  |  |
| group size | n | 3 | 2 | 1 |
| duration of first line treatment |  |  |  |  |
| total | mean | 61.3 | 30 | 100 |
| age <= 75 | mean | 21 | - | - |
| age > 75 | mean | 81.5 | 30 | 100 |
| HSCT yes a) | mean | - | - | - |
| HSCT no a) | mean | 61.3 | 30 | 100 |
| HSCT yes b) | mean | - | - | - |
| HSCT no b) | mean | 61.3 | 30 | 100 |
| time between treatments 1 and 2 | mean | 390 | 207 | - |
| time between treatments 1 and 2 or end of observation | mean | 532 | 207 | 31 |

**Table S3:** MM patients with hematopoietic stem cell transplantation (HSCT) according to year of HSCT. Analysis is based on 1054 prevalent patients from 2012 to 2017.

|  |  | Jahr 2012 | Jahr 2013 | Jahr 2014 | Jahr 2015 | Jahr 2016 | Jahr 2017 |
| --- | --- | --- | --- | --- | --- | --- | --- |
| **HSCT total** | n | 21 | 26 | 23 | 35 | 29 | 31 |
| **HSCT autologous** | n | 20 | 24 | 23 | 34 | 29 | 31 |
| age '<=75' | n (%) | 20 (100) | 24 (100) | 23 (100) | 34 (100) | 29 (100) | 31 (100) |
| age '>75' | n (%) | 0 ( 0) | 0 ( 0) | 0 ( 0) | 0 ( 0) | 0 ( 0) | 0 ( 0) |
| gender 'female' | n (%) | 10 (50) | 7 (29.2) | 14 (60.9) | 14 (41.2) | 16 (55.2) | 17 (54.8) |
| gender 'male' | n (%) | 10 (50) | 17 (70.8) | 9 (39.1) | 20 (58.8) | 13 (44.8) | 14 (45.2) |
| managed care coverage 'no' | n (%) | 7 (35) | 11 (45.8) | 13 (56.5) | 15 (44.1) | 14 (48.3) | 10 (32.3) |
| managed care coverage 'yes' | n (%) | 13 (65) | 13 (54.2) | 10 (43.5) | 19 (55.9) | 15 (51.7) | 21 (67.7) |
| all-diagnosis inpatient cost | mean (median) | 33'090 (29'931) | 31'306 (27'840) | 25'202 (19'372) | 30'489 (26'321) | 28'308 (27'222) | 24'597 (22'997) |
| **HSCT allogeneic** | n | 1 | 2 | 0 | 1 | 0 | 0 |

